# Molecular genetic influences on attentional control and other executive processes and their links with psychopathology in the AFFECT study

**DOI:** 10.1101/2025.02.14.25322257

**Authors:** Justin D. Tubbs, Travis T. Mallard, Maria Dalby, Yunxuan Jiang, Younga H. Lee, Karmel W. Choi, Tian Ge, Niels Plath, Lene Hammer-Helmich, Julie M. Granka, 23andMe Research Group, Andrew D. Grotzinger, David Hinds, Jordan W. Smoller, Joshua W. Buckholtz

## Abstract

**Background:** Attentional control is a critical component of executive functioning involved in numerous psychiatric and neurological disorders, yet its etiological relationships with many cognitive and behavioral phenotypes remain underexplored.

**Methods:** We conducted the first multivariate characterization of molecular genetic influences on attentional control and other executive processes in a cohort of more than 20,000 individuals enriched for mood disorders. We used Genomic Structural Equation Modeling to formally model patterns of genetic covariance among these task-based measures of cognition, as well as their relationships with other cognitive, clinical, and imaging-derived phenotypes.

**Results:** We identified two independent latent genetic factors: one broadly influencing executive function and one narrowly influencing attentional control. Both the *Common Executive Function (CEF)* and *Attentional Control (AC)* factors were genetically correlated with cognitive and clinical phenotypes, with each latent factor uniquely linked to liability for psychiatric disorders. For example, we observed myriad relationships between the factors and psychopathology, including robust and conditionally independent genetic associations with ADHD. However, despite clear links to brain-related phenotypes, genetic correlations with imaging-derived phenotypes themselves were modest and non-significant after correcting for multiple comparisons.

**Conclusions:** Overall, the results of our study suggest that genetic influences on attentional control are generally distinct from those that influence broader aspects of executive function. The *CEF* and *AC* factors show distinct patterns of genetic overlap with multiple cognitive and psychiatric outcomes, underscoring the need for more detailed phenotyping of cognition to generate new insights into the etiology of psychopathology.

## Introduction

Attentional control, a core component of executive functioning, refers to the ability to regulate and direct focus in line with task demands while resisting distractions. As part of the broader suite of executive processes, such as working memory and cognitive flexibility, attentional control is fundamental to higher-order cognition (1,2). It plays a critical role in daily functioning, impacting learning and memory, and is prominently implicated in numerous forms of psychiatric and neurological dysfunction. Consequently, attention has long been of interest to scientists and clinicians, as difficulties in this domain are prevalent across diverse patient populations, including those with psychiatric, neurodevelopmental, and neurodegenerative disorders, as well as acquired brain injuries (1,3–5). Underscoring its relevance to psychopathology, the National Institute of Mental Health has recognized multiple attentional processes as critical elements within the Research Domain Criteria framework (6).

Despite this keen interest, etiological influences on attentional control have not been well characterized. Decades of cognitive science and neuroscience have advanced our understanding of this mental process (1,2,7), but genetically informative studies have largely been limited to twin heritability studies (8–11) and relatively small genome-wide association studies (GWAS) (12–15). This slow progress is even more apparent when compared to GWAS of other complex traits related to cognition, such as general intelligence (16) and educational attainment (17), which have received considerably more focus and resources. Some components of executive functioning have even been the subject of multivariate genomic studies, which have advanced our understanding of shared etiology among mental processes (18). Given the substantial progress in understanding other aspects of cognition, characterizing etiological influences on attentional control could provide critical insights into the heritable components of an important biobehavioral phenotype.

In the present study, we address this gap in the literature by describing the largest GWAS of the gradual-onset continuous performance task (gradCPT), which measures the ability to maintain consistent responding to frequently presented stimuli for extended epochs and inhibit responding to infrequent, unpredictably presented stimuli. Specifically, we report GWAS results for multiple indices of attentional control measured by this task, as well as GWAS results for several broader executive processes in the AFFECT study (19), a large cohort enriched for patients with mood disorders (maximum *N*=23,318 individuals with European-like genomes). We then use Genomic Structural Equation Modeling (Genomic SEM) (20) to formally characterize patterns of genetic sharing and differentiation among these cognitive phenotypes, revealing two latent genetic factors that are largely unique from one another. Finally, to better understand the clinical and developmental impacts of attentional control, we evaluate genetic relationships between the latent factors and a variety of cognitive, neuropsychiatric, and imaging-derived phenotypes. Collectively, our results provide novel insights into the genetic architectures of higher-order cognitive processes and offer a more comprehensive understanding of their etiological landscape.

## Methods

### Sample recruitment and phenotyping procedures

Participants were recruited as part of the Affective disorders, Environment, and Cognitive Trait (AFFECT) study in collaboration with 23andMe, Inc (19). Study design, participant recruitment, and data collection are described in a prior publication (19). Briefly, participants were invited to participate in an online study involving genetic data collection and a series of surveys including task-based cognitive assessments over the course of 9 months. Recruitment was conducted using a combination of the 23andMe research participant pool and social media advertisements. The overall sample was designed to be enriched for participants with a self-reported mood disorder diagnosis (bipolar disorder [BD] or major depressive disorder [MDD]) with pharmacological treatment, while control participants were restricted to those reporting no history of major psychiatric diagnoses or prior psychiatric prescription. A total of 23,835 non-psychiatric controls, 14,768 MDD cases, and 9,864 BD cases completed consent procedures, provided a DNA sample, and answered the baseline set of questionnaires. Participants provided informed consent and volunteered to participate in the research online, under a protocol approved by the external AAHRPP-accredited IRB, Ethical & Independent (E&I) Review Services. As of 2022, E&I Review Services is part of Salus IRB (https://www.versiticlinicaltrials.org/salusirb).

An extensive cognitive battery was administered to each participant across eight online data collection waves (19). In the current study, we focus a subset of tasks from the battery that yield measures of distinct executive processes: the Digit-Symbol Substitution Test (DSST) (21), the gradCPT (22), and the THINC-Integrated Tool (THINC-it) (23), with the latter each being assessed at two separate assessments. From these tasks, a set of seven variables indexing conceptually distinct executive processes were selected for GWAS analysis. Processing speed was measured using the number of correct trials from the DSST. Measures of vigilance (sustained attention variability), performance monitoring, and response inhibition, were derived from the gradCPT: the coefficient of variation of response time for correct responses to target trials, post-error slowing (the difference in response time for target trials following correct vs. incorrect responses to targets), and commission errors, respectively. Measures of response selection, working memory, and cognitive flexibility derived from three of the THINC-it tasks: log-transformed mean reaction time for correct symbols in the “Spotter” task, number of correct responses in the “Symbol Check” task, and the time to complete the “Trails” task (similar to the Trails B neuropsychological test). If multiple observations were available for a participant the median value was used. All cognitive phenotypes were oriented such that positive values indicate better performance.

### Preprocessing, quality control, and GWAS procedures

Complete SNP genotyping and imputation procedures are described in a prior publication (19). Briefly, saliva samples underwent DNA extraction and genotyping at the National Genetics Institute, a CLIA-licensed clinical laboratory under the Laboratory Corporation of America. Genotyping, phasing, and imputation followed the standardized 23andMe pipeline (19). Following preprocessing and quality control, approximately 9.2 million high-quality genotyped and imputed SNPs were available for analysis.

All GWAS analyses were restricted to individuals with European-like genomes, as identified using a local ancestry-based algorithm (24) in a set of maximally unrelated individuals (25). GWAS were performed using linear regression on imputed and genotyped SNPs. Each cognitive phenotype was regressed on genotype dosage with sex, age, five genetic principal components, and genotyping platform as covariates. For each GWAS, SNPs with fewer than 20% of the available sample size for a given trait were removed prior to subsequent analyses. As noted in **Table 1**, final analytic sample sizes for each GWAS ranged from 10,129 participants with data on performance monitoring to 23,318 participants with data on processing speed.

**Table 1.** Cognitive Trait Heritability Summary.

| Assessment | Cognitive Measure | Summary | GWAS N | $h^2_{\text{SNP}}$ (SE) | $p$-value | Mean $\chi^2$ | Lambda GC | LDSC Intercept (SE) | Ratio (SE) |
| --- | --- | --- | --- | --- | --- | --- | --- | --- | --- |
| DSST | Correct trials | Processing Speed | 23,318 | 0.18 (0.03) | 3.4e-13 | 1.0927 | 1.0842 | 1.0041 (0.0078) | 0.0443 (0.0841) |
| gradCPT | Coefficient of Variation of reaction time in response to target trials | Vigilance (Variability in Sustained Attention over time) | 14,234 | 0.10 (0.04) | 5.9e-3 | 1.0336 | 1.0345 | 1.0042 (0.0071) | 0.1263 (0.2117) |
|  | Post-error slowing | Performance Monitoring | 10,129 | 0.12 (0.04) | 7.5e-3 | 1.0141 | 1.0119 | 0.9903 (0.0067) | -0.689 (0.4719) |
|  | Commission Errors | Response Inhibition | 14,234 | 0.18 (0.04) | 4.0e-5 | 1.0421 | 1.0386 | 0.9894 (0.0082) | -0.2525 (0.1959) |
| Thinc-IT | Reaction time for correct symbols in "Spotter" task | Response Selection | 18,653 | 0.15 (0.03) | 3.3e-9 | 1.0508 | 1.0506 | 0.9939 (0.0068) | -0.1195 (0.1341) |
|  | Number of correct responses in "Symbol Check" task | Working Memory | 18,192 | 0.10 (0.03) | 9.2e-5 | 1.0382 | 1.0446 | 1.0005 (0.0068) | 0.0124 (0.1779) |
|  | Time to complete "Trails" task | Cognitive Flexibility | 18,279 | 0.08 (0.03) | 4.3e-3 | 1.0388 | 1.0456 | 1.0111 (0.0072) | 0.2852 (0.1843) |

This table summarizes the results of LD score regression (LDSC) applied to genome-wide association study (GWAS) summary statistics for each cognitive trait. $h^2_{\text{SNP}}$ = SNP heritability. GC = genomic control.

GWAS summary statistics were prepared for downstream analysis using GenomicSEM R package version 0.0.5 (20). In line with best practices, variants were only retained if they had imputation quality scores > 0.9 (when available), minor allele frequency > 0.01, and were present in the HapMap3 reference panel. For external psychiatric phenotypes, the sum of effective sample sizes were calculated for use in GenomicSEM (26).

### Heritability, genetic correlation, and factor analyses

SNP-based heritability (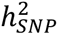) was estimated for each cognitive trait by applying LD score regression (LDSC) (27), as implemented in the GenomicSEM R package (20). Similarly, GenomicSEM was used to estimate the genome-wide covariance and sampling variance matrices via multivariable LDSC.

Two non-graphical scree tests were used to determine the optimal number of latent factors required to describe the observed genetic covariance matrix: the acceleration factor and optimal coordinates (28). Parallel analysis (29) was also performed as a complementary metric. These results led us to perform exploratory factor analysis (EFA) with promax rotation for one, two, and three latent factors (i.e., *k*±1) using the GenomicSEM R package. Model selection was guided by assessing incremental variance explained and parsimony of the implied latent factor structure.

Based on EFA model results, confirmatory factor analysis (CFA) was performed to assess the fit of the EFA-implied optimal factor model using unit variance identification. Model fit was evaluated using the model χ^2^ statistic, the comparative fit index (CFI), and the standardized root mean square residual (SRMR).

### Genetic correlations with other complex traits

Genetic correlations (*r*_g_) were estimated between the identified latent factors and other complex traits measured in external samples (**Supplementary Table 1**). These phenotypes were organized into three broad categories: cognition and education, psychopathology, and cortical morphology (see **Supplementary Table 1** for a complete list). A χ^2^ difference test was used to assess whether the latent factors differed in their genetic correlations with other traits. Essentially, this involved testing whether model fit significantly worsened when the *r*_g_ between each factor and the external trait were constrained to be equivalent Additionally, multiple genetic regression models were used to quantify the degree to which genetic associations between the factors and other traits were unique (i.e., the conditional genetic association). Finally, *Q*_Trait_ heterogeneity tests were used to evaluate the extent to which genetic relationships between the factors and an external trait were adequately explained by the common pathway implied by the factor structure (30). To account for multiple testing, the Benjamini-Hochberg procedure was used to control the false discovery rate within each family of tests for each factor.

## Results

### Attentional control and other executive processes are influenced by common variants

Univariate LDSC results for each of the seven executive processes are reported in **Table 1**, with Manhattan and QQ plots shown in **Supplementary Figure 1**. All phenotypes exhibited statistically significant 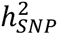, ranging from 0.08 for cognitive flexibility (SE=0.03) to 0.18 for both processing speed (SE=0.03) and response inhibition (SE=0.04). Despite exhibiting a nominally significant 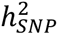, the THINC-it cognitive flexibility measure (time to completion on the “Trails” task) was excluded from subsequent analyses. Its inclusion required extensive smoothing to ensure a positive-definite genetic covariance matrix, suggesting that this GWAS had low statistical power and was unsuitable for factor analysis.

Genetic correlations among the six executive processes selected for analysis are presented in **Figure 1a** (**Supplementary Table 2**). Briefly, we observed two clusters of phenotypes. One cluster consisted of vigilance, response inhibition, and performance monitoring, which had modest to strong genetic correlations with one another, ranging from 0.21 to 0.77. The second cluster consisted of processing speed, working memory, and response selection, which showed relatively high positive genetic correlations, ranging from 0.58 to 0.83. These three traits exhibited strong negative genetic correlation with performance monitoring (-0.64, -0.35, -0.6, respectively).

**Figure 1.**
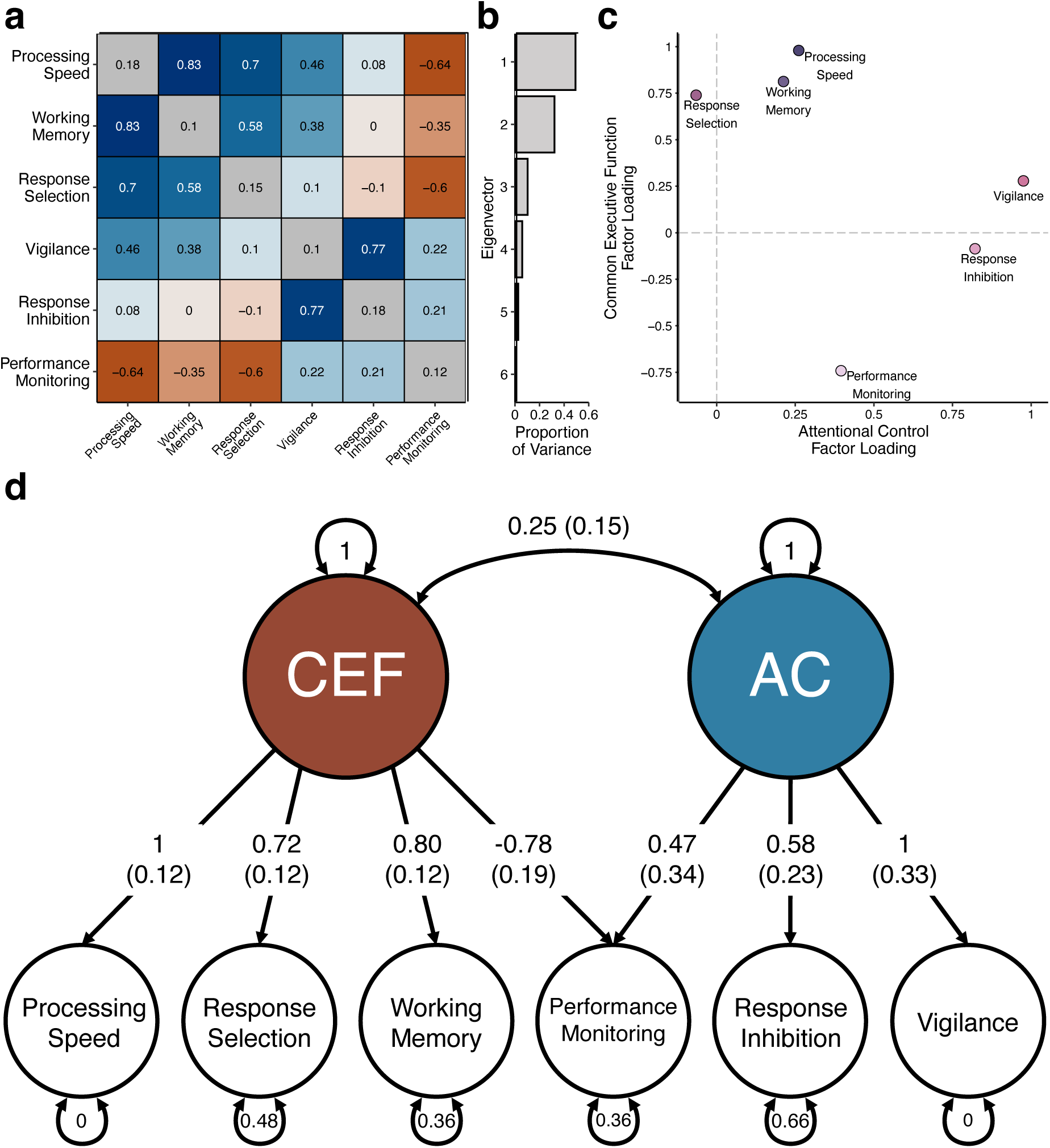
Genetic influences on common executive function and attentional control processes exhibit complex patterns of pleiotropy. **a,** A heatmap of the genetic correlations for the six cognitive traits with significant SNP heritabilities that were included in genomic factor analyses. Off-diagonal elements denote genetic correlation estimates, while diagonal elements correspond to the SNP heritabilities estimated by LD score regression. **b**, A barplot illustrating the proportion of variance explained by each eigenvector. **c,** The results of exploratory factor analysis plotted as a scatter plot, where the loading of each cognitive trait on F1 is plotted against its loading on the F2. **d,** A path diagram of the best fitting correlated factors model consisting of a *Common Executive Function* and *Attentional Control* factors. One-headed arrows reflect regression relationships, while two-headed arrows represent covariance relationships. A two-headed arrow connecting a variable to itself denotes (residual) variance. Dashed lines denote non-significant paths.

### Genomic factor analysis reveals complex patterns of pleiotropy among executive processes

Further examination of the standardized genetic covariance matrix revealed that the first two eigenvectors explained the majority (81.3%) of the total genetic variance across cognitive traits, accounting for 49.2% and 32.1% of the variance, respectively (**Figure 1b**). All three factor retention methods suggested that a two-factor solution would provide the best balance of fit and parsimony.

The results of our EFA confirmed that a two-factor solution provided a balance of parsimony, variance explained, and interpretability. While a single factor explained 46.6% of the total genetic variance, two correlated factors explained 78.3% of the variance. The addition of a third factor yielded marginal improvement, bringing the total variance explained to 84.1%, but contributed little in terms of interpretability, as the third factor largely explained variance in a single phenotype (performance monitoring). The results of our two-factor EFA are visualized in **Figure 1c** (**Supplementary Table 3**). Informed by these results, we used Genomic SEM to fit a confirmatory factor model with two latent genetic factors that were allowed to correlate (**Figure 1c**). Fit indices indicated that this model provided a reasonably good fit to the observed data (χ^2^_[7]_=17.07, CFI=0.96, SRMR=0.11). Although the residuals between observed and model-implied correlations were slightly above optimal thresholds, deviations were minor and not specific to any particular trait or combination of traits.

The first factor, which we refer to as *Common Executive Function (CEF)*, exhibited strong positive relationships with processing speed (λ=1, SE=0.12, *p*=4.42e-18), response selection (λ=0.72, SE=0.12, *p*=1.84e-9), and working memory (λ=0.80, SE=0.12, *p*=1.68e-10), as well as a strong negative relationship with performance monitoring (λ=-0.78, SE=0.19, *p*=4.58e-5). This pattern of factor loadings suggests that the *Common Executive Function* factor represents a general index of cognitive ability akin to the *g* factor. The second factor, which we refer to as *Attentional Control (AC)*, exhibited a strong positive relationship with vigilance (λ=1, SE=0.33, *p*=2.45e-3) and a moderate positive relationship with response inhibition (λ=0.58, SE=0.23, *p*=1.36e-2). Although performance monitoring was also allowed to load onto this factor per the EFA results, there was no statistically significant relationship (λ=0.47, SE=0.34, *p*=0.17). Thus, this latent factor appears to index processes more specifically related to the regulation of sustained attention rather than broader cognition per se. Interestingly, the modest correlation between the two latent genetic factors was not statistically significant (*r*_g_=0.25, SE=0.15, *p*=0.10), though this may be related to the lower statistical power of the *Attentional Control* factor.

### Attentional control has nuanced relationships with other aspects of cognition

To evaluate the validity of our latent factors, we first estimated genetic correlations between the modeled factors and cognitive traits measured in external samples. These results generally support our characterizations of the latent factors (**Figure 2a, Supplementary Table 4**). For example, *Common Executive Function* showed the strongest genetic correlations with a previously published GWAS of the *g* factor (*r*_g_=0.80, SE=0.07, *p*=1.4e-31) and an executive function factor (*r*_g_=0.80, SE=0.06, *p*=1.9e-42). In general, *Common Executive Function* exhibited stronger positive genetic correlations with each of the external cognitive traits than *Attentional Control*. Formal statistical tests indicated that these differences were statistically significant (*p*s≤8.8e-6, **Supplementary Table 5**). One notable exception to this pattern was educational attainment, where the moderately positive genetic correlations with *Common Executive Function* (*r*_g_=0.30, SE=0.04, *p*=4.7e-14) and *Attentional Control* (*r*_g_=0.26, SE=0.08, *p*=2.1e-3) were not significantly different from one another (*p*=0.07, **Supplementary Table 5**).

**Figure 2.**
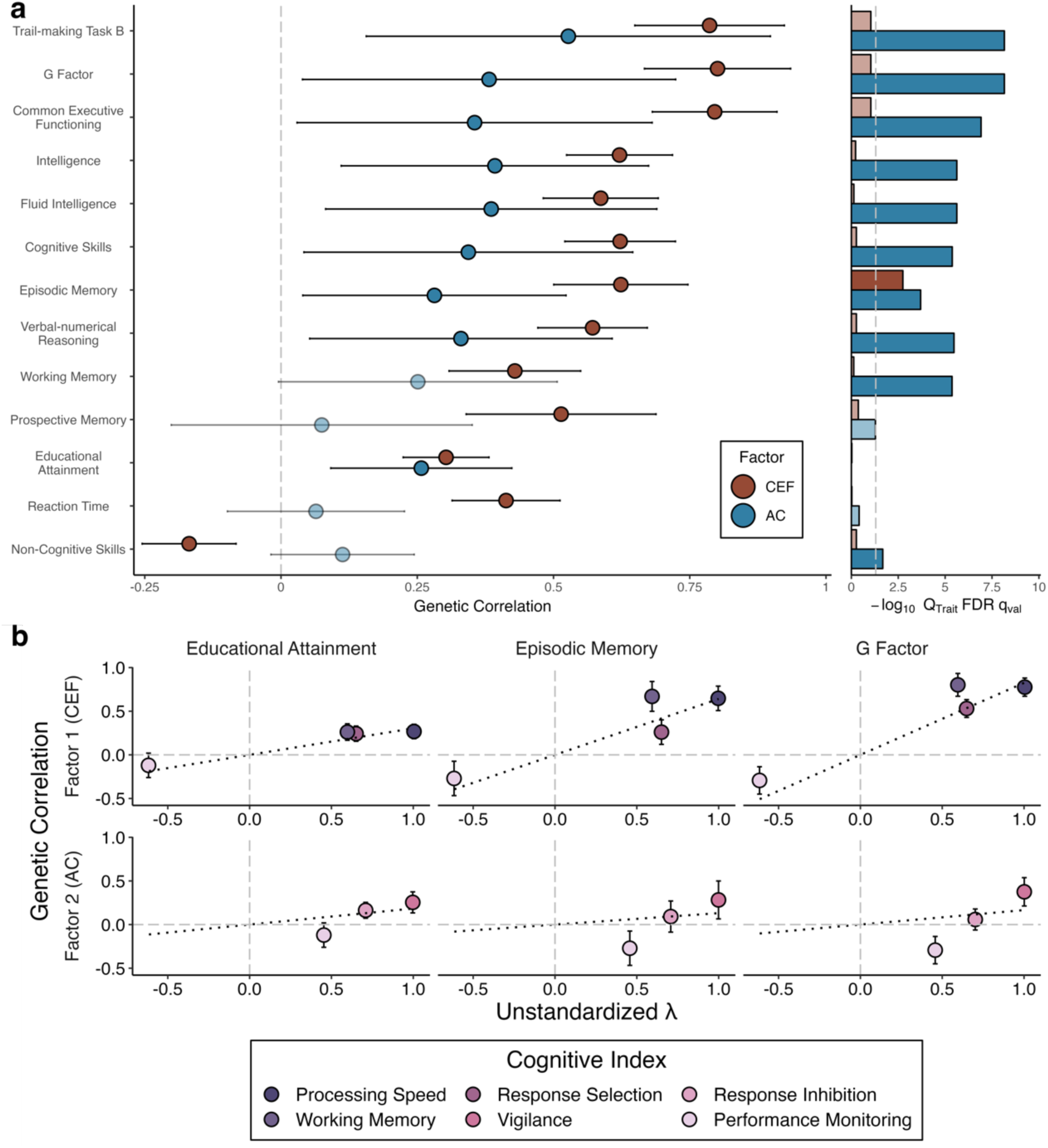
Latent cognitive factors differ in their genetic relationships with other cognitive phenotypes. **a, Scatter plot** summarizing the genetic relationships between cognitive phenotypes measured in other samples and the *Common Executive Function* (*CEF*) and *Attentional Control* (*AC*) factors. Note that a significant *Q*_Trait_ FDR *q*-value indicates that the genetic correlations with an external phenotype are inconsistent with the latent factor. In panel a, statistically significant genetic correlation estimates and *Q*_Trait_ FDR q-values are visualized with a solid fill, while non-significant estimates are visualized with a translucent fill. Statistical significance was determined by a false-discovery rate (FDR) adjusted *p*-value (q-value) below 0.05 using the Benjamini-Hochberg method within each factor across all external cognitive traits. **b,** Scatter plot granting further insight into *Q*_Trait_ results by plotting the genetic correlations between indicators and the external phenotype of interest as a function of the unstandardized factor loading (λ). Here, we present three exemplary external traits: educational attainment (non-significant *Q*_Trait_ for both factors), episodic memory (significant *Q*_Trait_ for both factors), and the *g* factor (significant *Q*_Trait_ for AC). Dotted lines show the least-squares line of best fit with the intercept fixed to zero. Deviations from this line reveal how genetic correlations between cognitive indicators and the external trait may be inconsistent with the model-implied structure.

To estimate the unique contributions of each latent factor to the observed associations with external cognitive traits, we jointly regressed each trait onto the *Common Executive Function* and *Attentional Control* factors (**Supplementary Table 6**). Despite most traits exhibiting at least a nominally significant genetic correlation with *Attentional Control*, these associations were notably attenuated when jointly regressed onto both factors simultaneously (**Supplementary Figures 2 & 3**). Only educational attainment (*p*=0.037) and non-cognitive skills (*p*=0.048) were significantly associated with *Attentional Control* when jointly regressed on *Common Executive Function*. Conversely, genetic associations between external cognitive traits and *Common Executive Function* did not exhibit significant attenuation. These results indicate that the slight, non-significant positive genetic correlation between the two latent factors may largely explain why external cognitive traits exhibit positive genetic correlations with *Attentional Control* – relationships that do not hold in conditional analyses.

Next, we performed *Q*_Trait_ analyses to evaluate heterogeneity in the patterns of association between each of the factor indicators and external cognitive phenotypes (**Supplementary Table 4**). When the relationship between an external trait and a latent factor has a significant *Q*_Trait_ *p*-value, there is evidence that the factor does not adequately explain the covariance patterns between the external trait and the factor’s indicators. Here, we found relatively low *Q*_Trait_ signal for the *Common Executive Function* factor, with only one episodic memory exhibiting significant heterogeneity (*Q*_Trait_ *p*=1.4e-3). However, statistically significant heterogeneity was observed for most of the significant genetic correlations between external cognitive phenotypes and the *Attentional Control factor*.

Patterns of heterogeneity are visualized in **Figure 2b** for the most significant *Q*_Trait_ signal (*p*=6.76e-10; *g* factor, *Attentional Control*), least significant *Q*_Trait_ signal (*p*1; educational attainment, *Attentional Control*), and episodic memory, which showed significant heterogeneity for both factors (*ps*=1.14e-4). *Q*_Trait_ results for the remaining external cognitive traits are displayed in **Supplementary Figure 4**. In general, we found that genetic correlations between external traits and *Common Executive Function* indicators varied as a function of the indicators’ unstandardized factor loadings (λ), suggesting that genetic relationships are consistent with the factor structure. However, this was often not the case for *Attentional Control* indicators, as performance monitoring often deviated from this pattern. Notably, performance monitoring was often estimated as having a negative genetic correlation with the external cognitive traits, but a positive loading on *Attentional Control*.

### Attentional control and common executive function have distinct links to psychopathology

To assess whether genetic influences on *Attentional Control* and *Common Executive Function* were related to risk for psychopathology, we estimated the genetic correlation between each latent factor and 13 psychiatric disorders measured in external samples (**Supplementary Table 1**). We found that most psychiatric disorders showed significant negative genetic correlations with *Common Executive Function*, while genetic correlations with *Attentional Control* tended to be significantly attenuated or not significantly different from zero (**Figure 3a, Supplementary Tables 4 & 5**).

**Figure 3.**
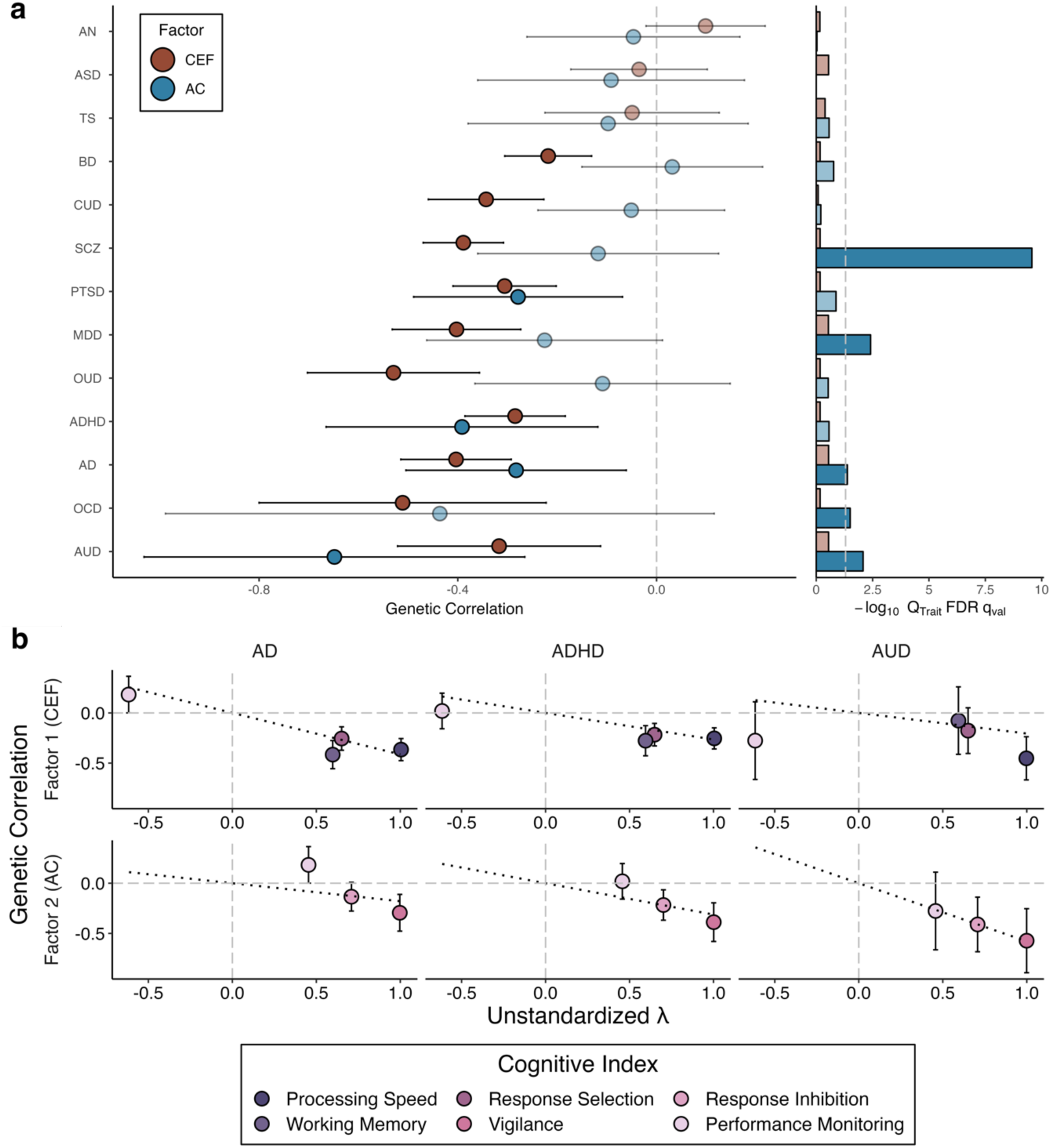
Latent cognitive factors differ in their genetic relationships with psychopathology. **a, Scatter plot** summarizing the genetic relationships between psychopathology and the *Common Executive Function* (*CEF*) and *Attentional Control* (*AC*) factors. Note that a significant *Q*_Trait_ FDR *p*-value indicates that the genetic correlations with an external phenotype are inconsistent with the latent factor. In panel **a**, statistically significant genetic correlation estimates and *Q*_Trait_ FDR q-values are visualized with a solid fill, while non-significant estimates are visualized with a translucent fill. Statistical significance was determined by a false-discovery rate (FDR) adjusted *p*-value (q-value) below 0.05 using the Benjamini-Hochberg method within each factor across all external psychiatric traits. **b,** Scatter plot granting further insight into *Q*_Trait_ results by plotting the genetic correlations between indicators and external phenotypes as a function of the unstandardized factor loading (λ). Here, we present three exemplary external traits: AD (significant *Q*_Trait_ for *AC*), ADHD (non-significant *Q*_Trait_ for both factors), and AUD (significant *Q*_Trait_ for *AC*). Dotted lines show the least-squares line of best fit with the intercept fixed to zero. Deviations from this line reveal how genetic correlations between cognitive indicators and the external trait may be inconsistent with the model-implied structure. **Note**: ADHD = attention-deficit hyperactivity disorder, AN = anorexia nervosa, AD = anxiety disorders, ASD = autism spectrum disorder, AUD = alcohol use disorder, BD = bipolar disorder, CUD = cannabis use disorder, MDD = major depressive disorder, OCD = obsessive compulsive disorder, OUD = opioid use disorder, PTSD = post-traumatic stress disorder, SCZ = schizophrenia, TS = Tourette’s syndrome.

After accounting for multiple testing, we found that *Common Executive Function* was significantly genetically correlated with ADHD (*r*_g_=-0.28, SE=0.05, *p*=3.67e-8), alcohol use disorder (*r*_g_=-0.31, SE=0.10, *p*=2.27e-3), anxiety disorder (*r*_g_=-0.40, SE=0.06, *p*=1.11e-12), bipolar disorder (*r*_g_=-0.22, SE=0.04, *p*=9.23e-7), cannabis use disorder (*r*_g_=-0.34, SE=0.06, *p*=6.87e-9), major depressive disorder (*r*_g_=-0.40, SE=0.07, *p*=8.95e-10), obsessive-compulsive disorder (*r*_g_=-0.51, SE=0.15, *p*=5.40e-4), opioid use disorder (*r*_g_=-0.53, SE=0.09, *p*=2.01e-9), posttraumatic stress disorder (*r*_g_=-0.31, SE=0.05, *p*=7.47e-9), and schizophrenia (*r*_g_=-0.38, SE=0.04, *p*=2.84e-21). *Attentional Control*, on the other hand, was only significantly genetically correlated with ADHD (*r*_g_=-0.39, SE=0.14, *p*=4.61e-3), alcohol use disorder (*r*_g_=-0.65, SE=0.20, *p*=9.15e-4), anxiety disorder (*r*_g_=-0.28, SE=0.11, *p*=1.20e-2), and posttraumatic stress disorder (*r*_g_=-0.28, SE=0.11, *p*=7.97e-3). Most of these significant genetic correlations were consistent with the factor structure, though alcohol use disorder and anxiety disorder did have significant *Q*_Trait_ signals for *Attentional Control* (**Figure 3b**).

The two latent factors exhibited significant differences in genetic overlap for several psychiatric disorders, suggesting that the reduced number of significant links between *Attentional Control* and psychopathology was not solely a consequence of lower statistical power (**Supplementary Table 5**). However, three of the four phenotypes that were significantly genetically correlated with both factors (ADHD, alcohol use disorder, and posttraumatic stress disorder) showed non-significant between-factor differences (*p*s≥0.16).

Given that both factors were significantly genetically correlated with these disorders, and those genetic correlations were statistically similar in magnitude, we next asked whether the genetic relationships were independent of one another. To test this, we fit genetic regression models that estimated the conditional genetic association between those psychiatric disorders and *Attentional Control* and *Common Executive Function*. Interestingly, we found that both factors were uniquely associated with ADHD (*Attentional Control: β*_g_=-0.34, SE=0.15, *p*=0.02*; Common Executive Function: β*_g_=-0.20, SE=0.08, *p*=5.82e-3 but only *Attentional Control* was uniquely associated with alcohol use disorder (*Attentional Control: β*_g_=-0.61, SE=0.21, *p*=3.47e-3*; Common Executive Function: β*_g_=-0.15, SE=0.13, *p*=0.24). Meanwhile, *Common Executive Function* was uniquely associated with anxiety disorder (*Attentional Control: β*_g_=-0.19, SE=0.13, *p*=0.14*; Common Executive Function: β*_g_=-0.35, SE=0.06, *p*=1.24e-8) and PTSD (*Attentional Control: β*_g_=-0.21, SE=0.11, *p*=0.055*; Common Executive Function: β*_g_=-0.25, SE=0.06, *p*=6.31e-6). These results suggest that the genetic overlap between ADHD and cognition is multifactorial in nature and independently related to diverse executive processes, while the genetic overlap between alcohol use disorder and cognition appears to be more circumscribed and related to attentional processes. On the other hand, genetic sharing between anxiety and cognition, as well as PTSD and cognition appear to be more closely related to facets of executive functioning, rather than attention.

### Genetic relationships between executive processes and cortical structure are modest

To investigate potential brain-behavior relationships, we estimated genetic correlations between each latent factor and 34 regional measures of cortical thickness and surface area (68 phenotypes total). After correcting for multiple comparisons, no statistically significant genetic correlations were found between either of the latent factors and cortical thickness or surface area measures (**Supplementary Table 4**). The patterning of these genetic relationships across the cortex are presented in **Figure 4a**. Although no robust genetic correlations were identified, we did observe nominally significant genetic correlations between the thickness of the pars opercularis and *Common Executive Function* (*r*_g_=0.22, SE=0.07, *p*=0.003), as well as thickness in the precentral region and *Attentional Control* (*r*_g_=0.34, SE=0.15, *p*=0.03). Likewise, five regions showed nominally significant genetic correlation between surface area and *Common Executive Function*: the pars triangularis (*r*_g_=0.20, SE=0.07, p=0.005), transverse temporal (*r*_g_=0.17, SE=0.07, p=0.02), lateral orbitofrontal (*r*_g_=0.14, SE=0.06, p=0.03), medial orbitofrontal (*r*_g_=0.16, SE=0.07, p=0.03), and pars orbitalis (*r*_g_=0.14, SE=0.07, p=0.04). *Q*_Trait_ results indicated there was relatively little indicator-level heterogeneity for these relationships (**Supplementary Table 4, Supplementary Figure 4**).

**Figure 4.**
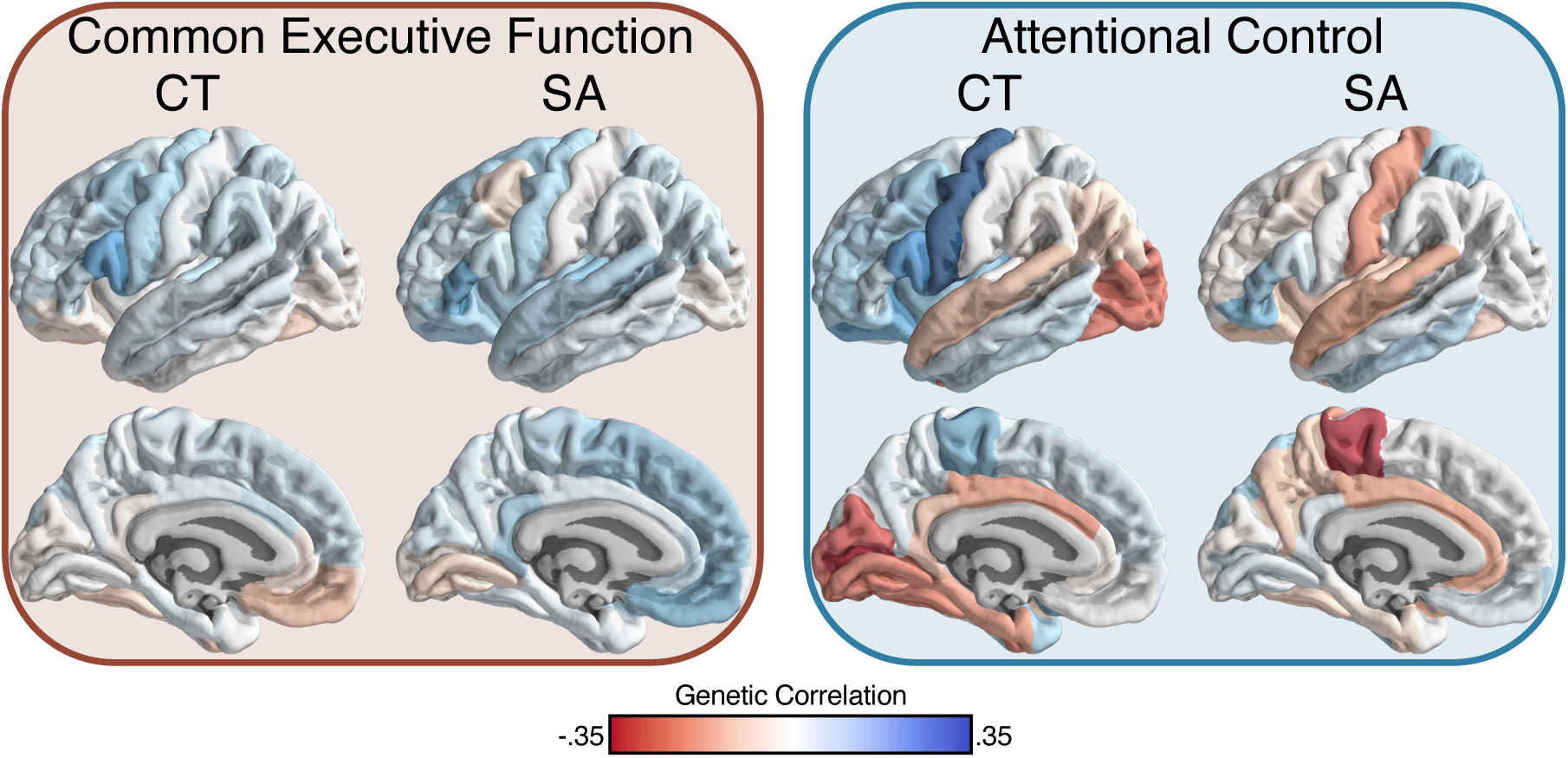
Genetic correlations between latent cognitive factors and cortical morphometry are modest and diffuse. Cortical map of the genetic relationships between the *Common Executive Function* (*CEF*) and *Attentional Control* (*AC*) factors and cortical thickness (CT) and surface area (SA). Values plotted represent the genetic correlation between a given factor and a given regional image-derived phenotype.

## Discussion

Attentional control is a fundamental aspect of higher-order cognition that is integral to learning, memory, and overall cognitive health. Using data from the AFFECT study, we report the first multivariate examination of molecular genetic influences on multiple attentional control processes, providing novel insights into their complex genetic architecture. Our findings reveal that attentional control is significantly heritable and influenced by common genetic variants with complex patterns of pleiotropy. These influences exhibit modest patterns of genetic overlap with other cognitive phenotypes and unique patterns of genetic association with psychopathology independent of executive functioning.

The results of our study provide the first large-scale empirical evidence that individual differences in attentional control processes are influenced by a shared polygenic common variant architecture. Consistent with findings from prior GWAS of related traits, SNP-based heritability estimates for the cognitive phenotypes considered here were generally modest for task-based measures of cognitive and attentional abilities, ranging from 0.08 for cognitive flexibility to 0.18 for processing speed and response inhibition. Moreover, the common genetic variants influencing these phenotypes tended to be quite pleiotropic, but with fairly distinct patterns of genetic sharing observed across the six phenotypes. We found that patterns of genetic sharing tended to segregate across distinct cognitive domains, with processing speed, working memory, and response selection being strongly genetically correlated with each other, while vigilance, response inhibition, and performance monitoring were more genetically interrelated. Our findings are generally in line with prior phenotypic factor analyses, which found that measures derived from the gradCPT tend to be distinct from other executive functioning processes, and even measures of general attentional capacity (31). Vigilance (i.e., sustained attention), which loaded most strongly onto the *Attentional Control* factor, also appears to exhibit life-course trajectories distinct from those related to fluid intelligence components of executive functioning (32).

Our genomic factor analysis provided deeper insights into the observed patterns of complex genetic sharing and differentiation, underscoring the segregation of genetic covariance into two distinct domains. We found that the observed patterns were best explained by two latent genetic factors: one that broadly influences executive functioning, akin to Spearman’s *g* (33), and one that more narrowly influences attentional control. Interestingly, the *Common Executive Function* and *Attentional Control* factors were largely unique of one another. Our genetic correlation analyses substantiated the convergent and discriminant validity of these factors, as both were significantly related to a wide variety of cognitive traits, with *Common Executive Function* exhibiting stronger and broader patterns of associations, as expected. Although associations between *Attentional Control* and other cognitive phenotypes could largely be attributed to its modest genetic overlap with *Common Executive Function*, both factors exhibited significant independent associations with educational attainment. In line with these independent genetic contributions, structural and functional neuroimaging studies suggest these cognitive processes involve fairly distinct neural pathways. While frontal and parietal regions have been most strongly linked to executive functioning task performance (34,35), sustained attention processes have been linked to a wider range of brain regions including those in visual, somatomotor, and insular cortices along with subcortical areas (36,37).

When we examined genomic relationships between the *Common Executive Function* and *Attentional Control* factors and psychiatric outcomes, we found that both factors were generally negatively correlated with clinical phenotypes, with stronger relationships observed for the *Common Executive Function* factor. After correcting for multiple, we found that *Attentional Control* was only significantly genetically correlated with ADHD, alcohol use disorder, anxiety disorder, and posttraumatic stress disorder, while *Common Executive Function* was significantly genetically correlated with ADHD, anxiety disorder, bipolar disorder, cannabis use disorder, major depressive disorder, and schizophrenia. This difference is likely attributable, in part, to the narrower focus of attentional control as a cognitive construct. Interestingly, follow-up genetic regression models revealed that *Attentional Control* and *Common Executive Function* are each uniquely genetically related to ADHD, indicative of a multi-faceted etiological relationship between cognition and ADHD. These results highlight the importance of considering a more comprehensive range of cognitive domains beyond executive functioning when studying cognitive differences across disorders.

Finally, despite clear links to brain-related traits and outcomes, we found that the *Common Executive Function* and *Attentional Control* factors were not robustly genetically correlated with common measures of brain structure themselves. After correcting for multiple comparisons, we found no significant genetic correlations between either factor and cortical thickness or surface area. Given the modest effect sizes of brain-behavior relationships at both phenotypic and genetic levels of analysis (38–40), it is likely that the executive process GWAS reported here are not sufficiently well-powered to characterize nuanced etiological relationships in relatively small imaging genetic samples.

Several limitations should be considered when interpreting the present findings. Most notably, although our GWAS of task-based attentional processes are among the largest to date, the AFFECT study sample is still considerably smaller than recently published molecular genetic studies of other executive functions (for example, those in the UK Biobank). While the present study is generally well-powered for characterizing the genome-wide correlational structure of executive processes and investigating their links to other complex traits, it is not well-powered for variant discovery. This is especially true given the modest SNP heritability estimates of investigated traits. Furthermore, in order to reduce confounding related to population stratification, the current analyses were restricted to those of European ancestry. As AFFECT study samples were collected in collaboration with a consumer genetics company, it is also possible that there are unique ascertainment biases present. Finally, this cohort is enriched for patients with mood disorders, which may limit the generalizability of its findings to non-psychiatric populations.

Nevertheless, our study describes the first multivariate genomic examination of attentional control and its links to other cognitive and psychiatric phenotypes. We found that attentional control processes are influenced by genetic factors that are relatively distinct from those that influence general executive functioning. Critically, we identified several instances of unique or differential genetic overlap with psychiatric disorders like ADHD, which indicates that psychopathology-associated cognitive changes are multifactorial in nature. We also found evidence of unique genetic relationships with educational attainment, where *Attentional Control* and *Common Executive Function* were jointly related to this complex life outcome. Our results suggest that expanding the scope of cognitive traits studied may lead to additional insights into the genetic architecture of specific cognitive processes and their relationships with mental health.

## Data Availability Statement

The full GWAS summary statistics for the 23andMe discovery data set will be made available through 23andMe to qualified researchers under an agreement with 23andMe that protects the privacy of the 23andMe participants. Datasets will be made available at no cost for academic use. Please visit https://research.23andme.com/collaborate/#dataset-access/ for more information and to apply to access the data.

## Acknowledgments

We would like to thank the research participants and employees of 23andMe for making this work possible.

The following members of the 23andMe Research Team contributed to this study: Stella Aslibekyan, Adam Auton, Elizabeth Babalola, Robert K. Bell, Jessica Bielenberg, Ninad S. Chaudhary, Zayn Cochinwala, Sayantan Das, Emily DelloRusso, Payam Dibaeinia, Sarah L. Elson, Nicholas Eriksson, Chris Eijsbouts, Teresa Filshtein, Pierre Fontanillas, Davide Foletti, Will Freyman, Zach Fuller, Julie M. Granka, Chris German, Éadaoin Harney, Alejandro Hernandez, Barry Hicks, Michael V. Holmes, M. Reza Jabalameli, Ethan M. Jewett, Yunxuan Jiang, Sotiris Karagounis, Lucy Kaufmann, Matt Kmiecik, Katelyn Kukar, Alan Kwong, Keng-Han Lin, Yanyu Liang, Bianca A. Llamas, Aly Khan, Steven J. Micheletti, Matthew H. McIntyre, Meghan E. Moreno, Priyanka Nandakumar, Dominique T. Nguyen, Jared O’Connell, Steve Pitts, G. David Poznik, Alexandra Reynoso, Shubham Saini, Morgan Schumacher, Leah Selcer, Anjali J. Shastri, Jingchunzi Shi, Suyash Shringarpure, Keaton Stagaman, Teague Sterling, Qiaojuan Jane Su, Joyce Y. Tung, Susana A. Tat, Vinh Tran, Xin Wang, Wei Wang, Catherine H. Weldon, Amy L. Williams, Peter Wilton

We would also like to thank Sharin Fuller at 23andMe for dedicated project support.

J.D.T. was supported by the Mass General Brigham Training Program in Precision and Genomic Medicine (T32HG010464). T.T.M. is supported by NIH grant K08MH135343. The AFFECT study was funded by H. Lundbeck A/S and the Milken Institute.

## Conflicts of Interest

Y.J., J.M.G., and D.H. are employed by and hold stock or stock options in 23andMe, Inc. M.D. and N.P. are employees of MUNA Therapeutics. L.H-H. is an employee of H. Lundbeck A/S. J.W.S. is a member of the Scientific Advisory Board of Sensorium Therapeutics (with equity) and has received an honorarium for an internal seminar Tempus Labs.

## Supplementary Information

**Supplementary Table 1.** This table lists the external trait GWAS sample sizes and references.

**Supplementary Table 2.** This table provides the genetic correlations (*r*_g_), their standard errors (SEs) and associated *p*-values for each pair of cognitive traits assessed in this study, estimated using LD score regression.

**Supplementary Table 3.** This table provides factor loadings for each cognitive indicator from exploratory factor analysis with a two-factor solution.

**Supplementary Table 4.** This table provides the genetic correlations (*r*_g_), their standard errors (SEs), associated *p*-values, and *Q*_Trait_ *p*-values for each external trait with the *Common Executive Function* and *Attentional Control* factors.

**Supplementary Table 5.** This table lists the results of a test for the difference in genetic correlations with external traits between the *Common Executive Function* and *Attentional Control* factors. Specifically, we test for a difference in model χ^2^ statistics between a model where the genetic correlation with a given external trait and each factor are freely estimated (χ^2^ Free) and a model where the trait-factor genetic correlations are constrained to be equivalent (χ^2^ Constrained). *P*-values are calculated as the area above the critical value (χ^2^ Diff = χ^2^ Free - χ^2^ Constrained) of a χ^2^ distribution with one degree of freedom. For some traits, the constrained model was unable to converge, and the corresponding columns are omitted.

**Supplementary Table 6.** This table provides the regression coefficients (*β*), their standard errors (SEs) and associated *p*-values for each external trait on the *Common Executive Function* and *Attentional Control* factors, where both factors are modeled simultaneously.

**Supplementary Figure 1.**
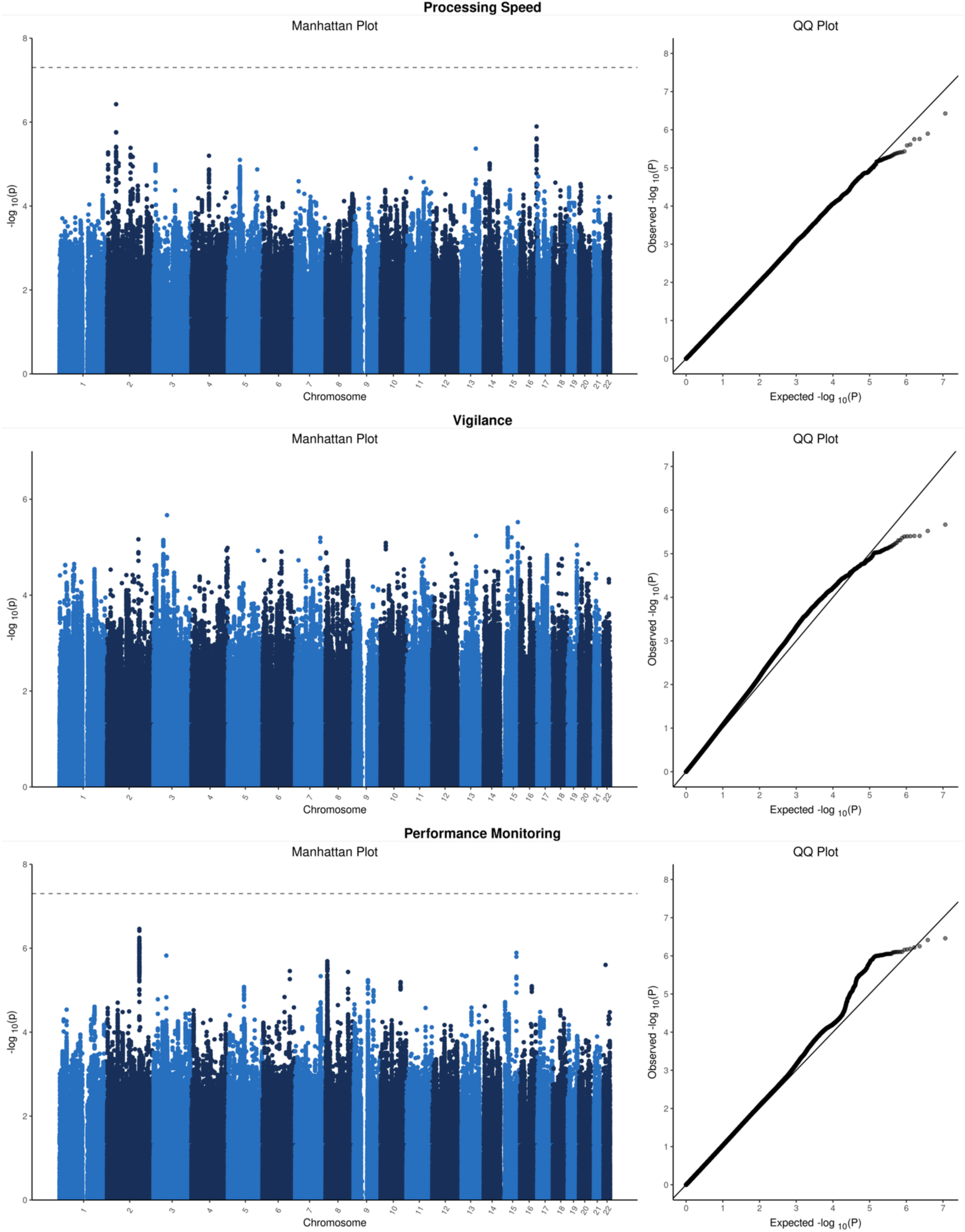
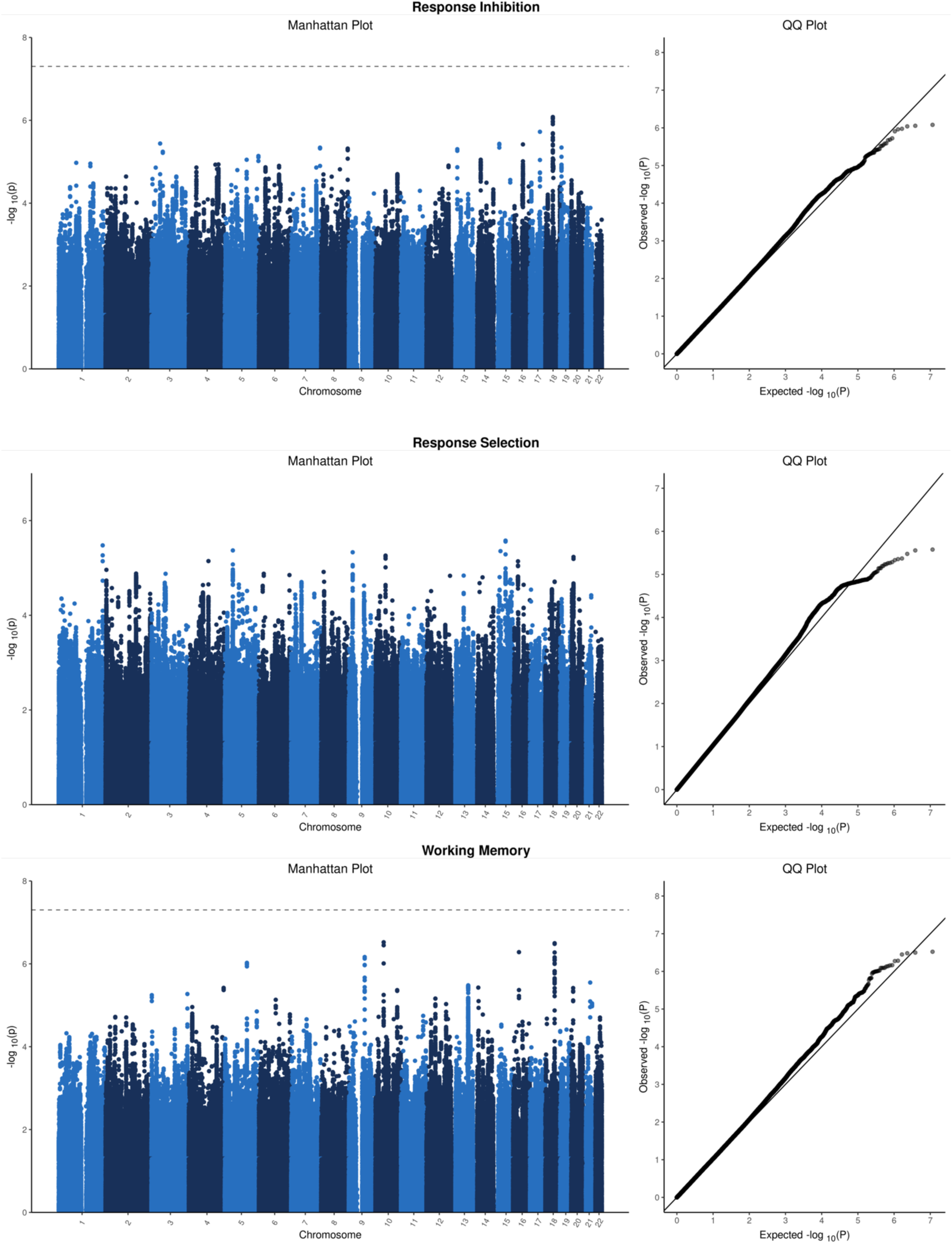
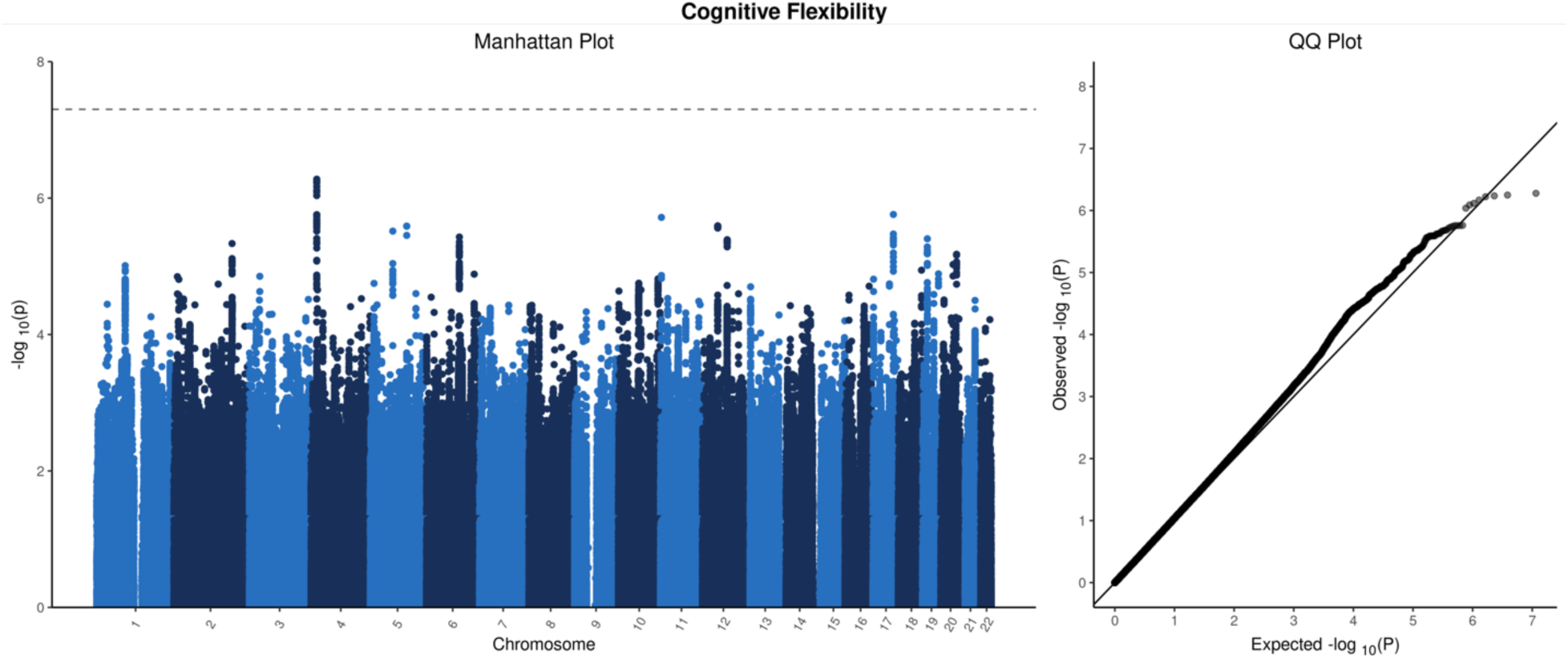
Manhattan and Q-Q Plots for seven cognitive phenotypes. This figure plots the results from genome-wide association studies (GWAS) for each of the seven cognitive phenotypes examined. Manhattan plots show the -log10(*p*-value) of each variant against their physical genomic position. Q-Q plots compare the ordered observed -log10(*p*-values) versus the null expectation of uniformly distributed *p*-values.

**Supplementary Figure 1.**
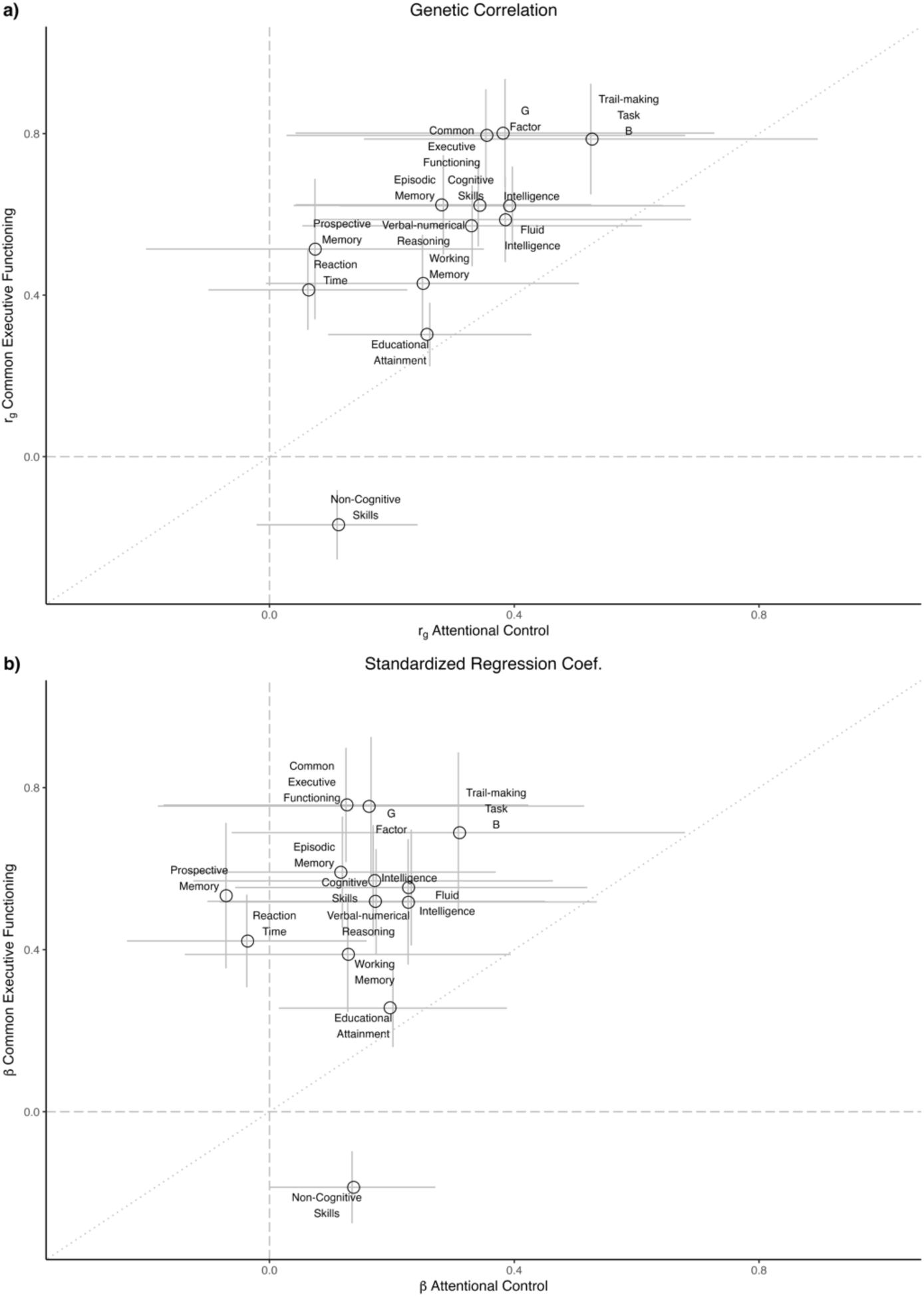
Cross-factor comparison of genetic correlations and Standardized regression coefficients with external cognitive traits. Panel **(a)** plots the genetic correlation (*r*_g_) between each external cognitive trait and the *Common Executive Function* (CEF) factor versus the corresponding *r*_g_ with *Attentional Control* (AC) factor. Panel **(b)** plots the corresponding standardized regression coefficients (*β*) for each trait on AC versus CEF when both are modeled simultaneously. The dotted line has an intercept of zero and slope of one, which would indicate perfect concordance between the estimated genetic correlation or regression of each trait with AC and CEF. Genetic correlations appear to be systematically higher across traits for CEF compared to AC. Most regression coefficients of cognitive traits on AC are not significantly different from zero while simultaneously modeling CEF.

**Supplementary Figure 2.**
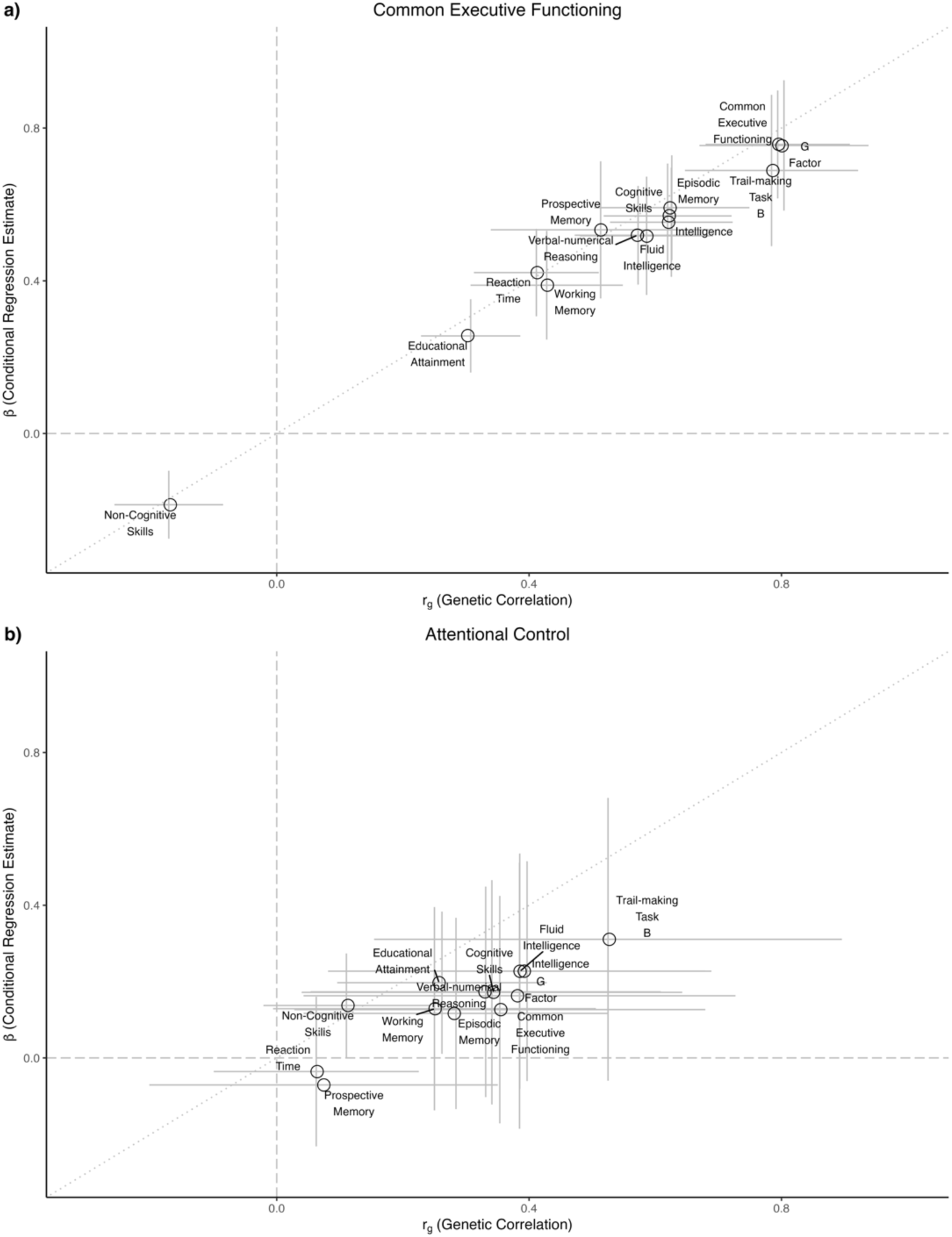
Within-factor comparison of genetic correlations and standardized regression coefficients with external cognitive traits. Panel **(a)** plots the genetic correlation (*r*_g_) between each external cognitive trait and the *Common Executive* Function (CEF) factor versus the corresponding standardized regression coefficient (*β*) of the trait on CEF. Panel **(b)** plots the *r*_g_ between each external cognitive trait and the *Attentional Control* (AC) factor versus the corresponding standardized regression coefficient (*β*) of the trait on AC. The dotted line has an intercept of zero and slope of one, which would indicate perfect concordance between the genetic correlation and regression estimates. While most points fall on the line for CEF, AC points appear to be attenuated such that regression coefficients are systematically lower than genetic correlation estimates.

**Supplementary Figure 3.**
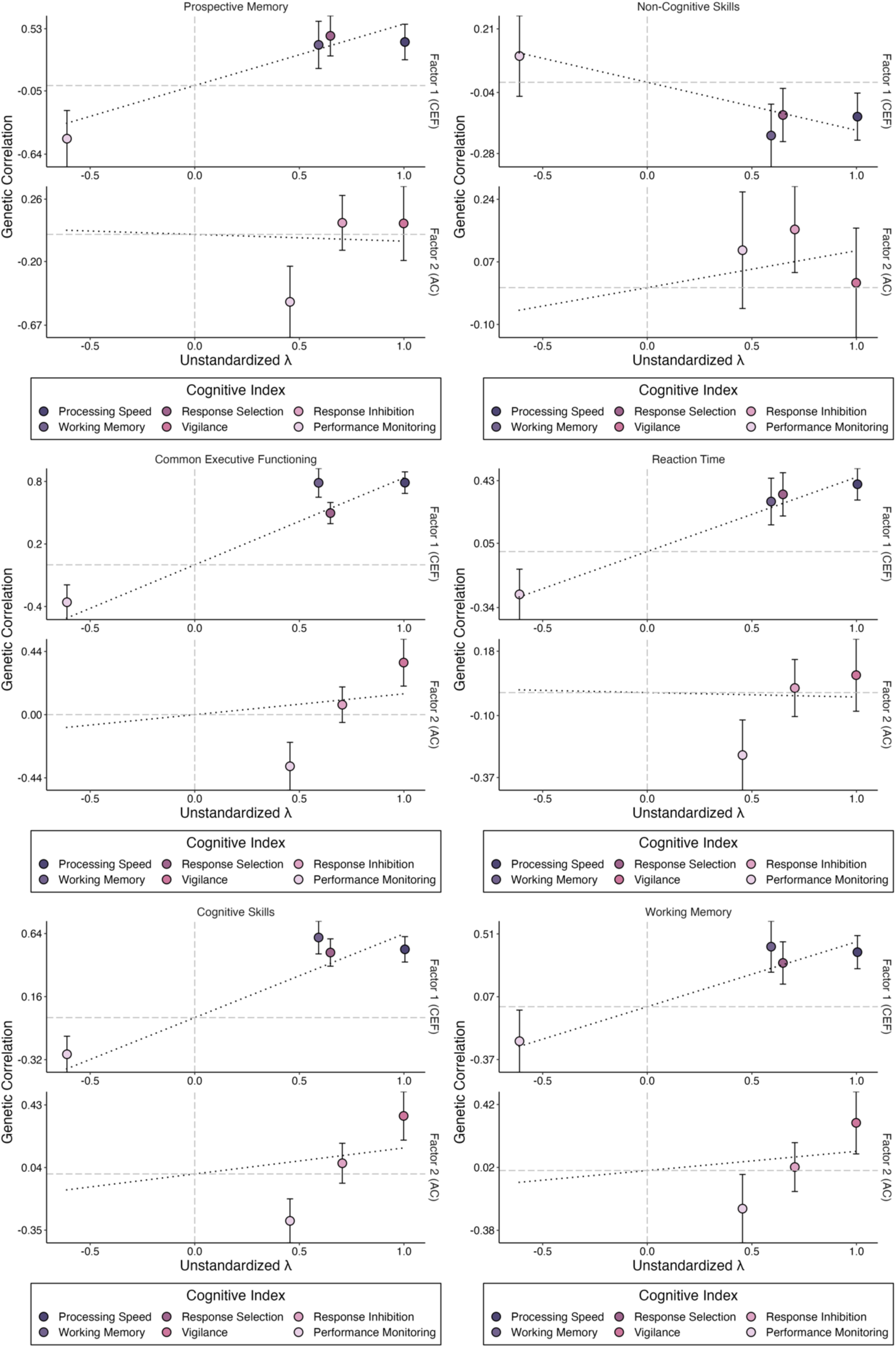
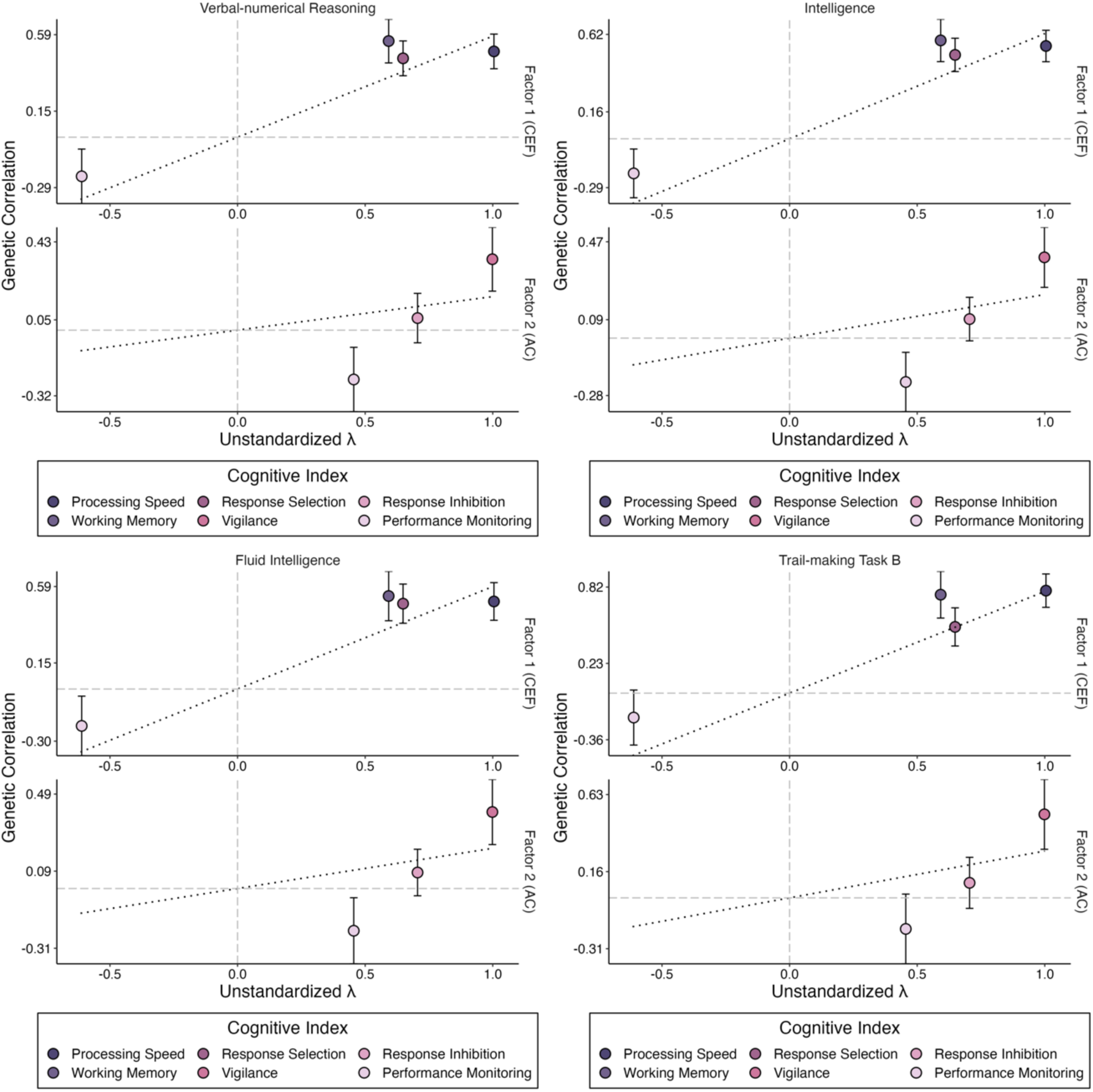
External cognitive trait heterogeneity plots. These figures plot the relationship between the loading of each indicator onto each latent factor (unstandardized λ) and the bivariate genetic correlation between each indicator and the external cognitive trait of interest. Dotted lines show the least-squares line of best fit, while constraining the intercept to be zero. Deviation from this line indicates cognitive indicators which are expected to have associations with external traits inconsistent with the estimated latent factor model.

**Supplementary Figure 4.**
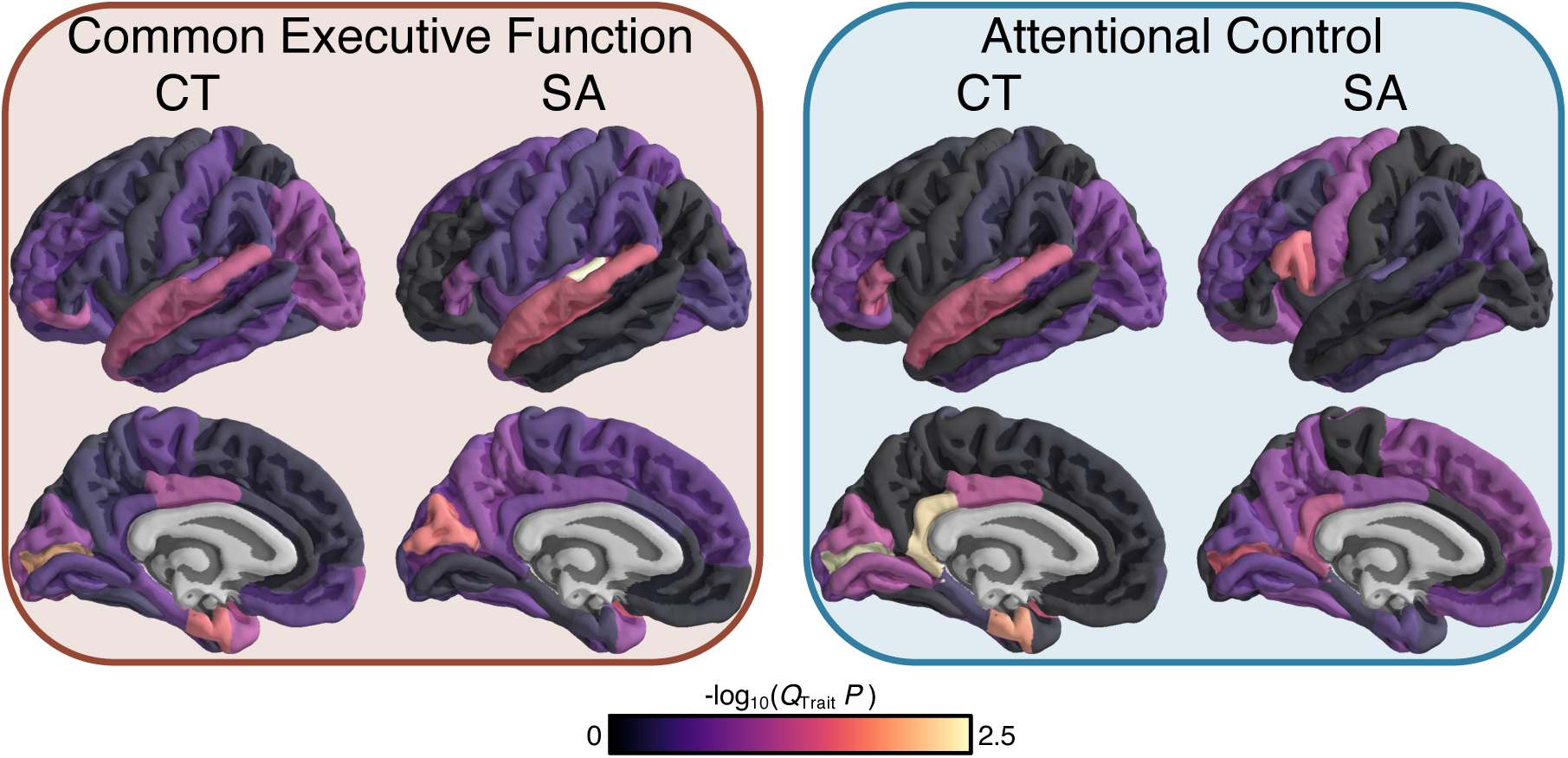
Q_Trait_ heterogeneity across cortical measures. Cortical map of the genetic heterogeneity between the *Common Executive Function* (*CEF*) and *Attentional Control* (*AC*) factors and cortical thickness (CT) and surface area (SA). Values plotted represent the - log(Q_Trait_ p-value) testing for significant heterogeneity between a given factor and a given regional image-derived phenotype.

## Notes

### Author Declarations

Ethical & Independent (E&I) Review Services gave ethical approval for this work.

## References

1. Fortenbaugh FC, DeGutis J, Esterman M (2017): Recent theoretical, neural, and clinical advances in sustained attention research. Ann N Y Acad Sci 1396: 70–91.

2. Esterman M, Rothlein D (2019): Models of sustained attention. Curr Opin Psychol 29: 174– 180.

3. Huang-Pollock CL, Karalunas SL, Tam H, Moore AN (2012): Evaluating vigilance deficits in ADHD: a meta-analysis of CPT performance. J Abnorm Psychol 121: 360–371.

4. Abramovitch A, Abramowitz JS, Mittelman A (2013): The neuropsychology of adult obsessive-compulsive disorder: a meta-analysis. Clin Psychol Rev 33: 1163–1171.

5. Little B, Anwyll M, Norsworthy L, Corbett L, Schultz-Froggatt M, Gallagher P (2024): Processing speed and sustained attention in bipolar disorder and major depressive disorder: A systematic review and meta-analysis. Bipolar Disord 26: 109–128.

6. Insel T, Cuthbert B, Garvey M, Heinssen R, Pine DS, Quinn K, et al. (2010): Research domain criteria (RDoC): toward a new classification framework for research on mental disorders. Am J Psychiatry 167: 748–751.

7. Cieslik EC, Mueller VI, Eickhoff CR, Langner R, Eickhoff SB (2015): Three key regions for supervisory attentional control: evidence from neuroimaging meta-analyses. Neurosci Biobehav Rev 48: 22–34.

8. Groot AS, de Sonneville LMJ, Stins JF, Boomsma DI (2004): Familial influences on sustained attention and inhibition in preschoolers. J Child Psychol Psychiatry 45: 306–314.

9. Holmes J, Hever T, Hewitt L, Ball C, Taylor E, Rubia K, Thapar A (2002): A pilot twin study of psychological measures of attention deficit hyperactivity disorder. Behav Genet 32: 389–395.

10. Gagne JR, O’Sullivan DL, Schmidt NL, Spann CA, Goldsmith HH (2017): The shared etiology of attentional control and anxiety: An adolescent twin study. J Res Adolesc 27: 122–138.

11. Myles-Worsley M, Coon H (1997): Genetic and developmental factors in spontaneous selective attention: a study of normal twins. Psychiatry Res 71: 163–174.

12. Alemany S, Vilor-Tejedor N, Bustamante M, Pujol J, Macià D, Martínez-Vilavella G, et al. (2016): A Genome-wide Association Study of attention function in a population-based sample of children. PLoS One 11: e0163048.

13. Arnatkeviciute A, Lemire M, Morrison C, Mooney M, Ryabinin P, Roslin NM, et al. (2023): Trans-ancestry meta-analysis of genome wide association studies of inhibitory control. Mol Psychiatry 28: 4175–4184.

14. Hatzimanolis A, Bhatnagar P, Moes A, Wang R, Roussos P, Bitsios P, et al. (2015): Common genetic variation and schizophrenia polygenic risk influence neurocognitive performance in young adulthood. Am J Med Genet B Neuropsychiatr Genet 168B: 392–401.

15. Liu H, Zhao X, Xue G, Chen C, Dong Q, Gao X, et al. (2023): TTLL11 gene is associated with sustained attention performance and brain networks: A genome-wide association study of a healthy Chinese sample. Genes Brain Behav 22: e12835.

16. Savage JE, Jansen PR, Stringer S, Watanabe K, Bryois J, de Leeuw CA, et al. (2018): Genome-wide association meta-analysis in 269,867 individuals identifies new genetic and functional links to intelligence. Nat Genet 50: 912–919.

17. Okbay A, Wu Y, Wang N, Jayashankar H, Bennett M, Nehzati SM, et al. (2022): Polygenic prediction of educational attainment within and between families from genome-wide association analyses in 3 million individuals. Nat Genet 54: 437–449.

18. de la Fuente J, Davies G, Grotzinger AD, Tucker-Drob EM, Deary IJ (2021): A general dimension of genetic sharing across diverse cognitive traits inferred from molecular data. Nat Hum Behav 5: 49–58.

19. Dalby M, Vitezic M, Plath N, Hammer-Helmich L, Jiang Y, Tian C, et al. (2022): Characterizing mood disorders in the AFFECT study: a large, longitudinal, and phenotypically rich genetic cohort in the US. Transl Psychiatry 12: 121.

20. Grotzinger AD, Rhemtulla M, de Vlaming R, Ritchie SJ, Mallard TT, Hill WD, et al. (2019): Genomic structural equation modelling provides insights into the multivariate genetic architecture of complex traits. Nat Hum Behav 3: 513–525.

21. Lezak MD (1995): Neuropsychological Assessment*. 3rd Ed*. Oxford University Press, p 24.

22. Rosenberg M, Noonan S, DeGutis J, Esterman M (2013): Sustaining visual attention in the face of distraction: a novel gradual-onset continuous performance task. Atten Percept Psychophys 75: 426–439.

23. McIntyre RS, Best MW, Bowie CR, Carmona NE, Cha DS, Lee Y, et al. (2017): The THINC-Integrated Tool (THINC-it) Screening Assessment for Cognitive Dysfunction: Validation in Patients With Major Depressive Disorder. J Clin Psychiatry 78: 873–881.

24. Durand EY, Do CB, Mountain JL, Macpherson JM (2014, October 18): Ancestry Composition: A novel, efficient pipeline for ancestry deconvolution. BioRxiv. bioRxiv, p 010512.

25. Henn BM, Hon L, Macpherson JM, Eriksson N, Saxonov S, Pe’er I, Mountain JL (2012): Cryptic distant relatives are common in both isolated and cosmopolitan genetic samples. PLoS One 7: e34267.

26. Grotzinger AD, Fuente J de la, Privé F, Nivard MG, Tucker-Drob EM (2023): Pervasive downward bias in estimates of liability-scale heritability in genome-wide association study meta-analysis: A simple solution. Biol Psychiatry 93: 29–36.

27. Bulik-Sullivan B, Loh P-R, Finucane HK, Ripke S, Yang J, Schizophrenia Working Group of the Psychiatric Genomics Consortium, et al. (2015): LD Score regression distinguishes confounding from polygenicity in genome-wide association studies. Nat Genet 47: 291–295.

28. Raîche G, Walls TA, Magis D, Riopel M, Blais J-G (2013): Non-graphical solutions for Cattell’s scree test. Methodology 9: 23–29.

29. Horn JL (1965): A Rationale and Test for the Number of Factors in Factor Analysis. Psychometrika 30: 179–185.

30. Grotzinger AD, Mallard TT, Akingbuwa WA, Ip HF, Adams MJ, Lewis CM, et al. (2022): Genetic architecture of 11 major psychiatric disorders at biobehavioral, functional genomic and molecular genetic levels of analysis. Nat Genet 54: 548–559.

31. Treviño M, Zhu X, Lu YY, Scheuer LS, Passell E, Huang GC, et al. (2021): How do we measure attention? Using factor analysis to establish construct validity of neuropsychological tests. *Cogn Res Princ Implic* 6: 51.

32. Fortenbaugh FC, DeGutis J, Germine L, Wilmer JB, Grosso M, Russo K, Esterman M (2015): Sustained Attention Across the Life Span in a Sample of 10,000: Dissociating Ability and Strategy. Psychol Sci 26: 1497–1510.

33. Spearman C (1904): “General Intelligence,” Objectively Determined and Measured. Am J Psychol 15: 201–292.

34. Nowrangi MA, Lyketsos C, Rao V, Munro CA (2014): Systematic review of neuroimaging correlates of executive functioning: converging evidence from different clinical populations. J Neuropsychiatry Clin Neurosci 26: 114–125.

35. Yuan P, Raz N (2014): Prefrontal cortex and executive functions in healthy adults: a meta-analysis of structural neuroimaging studies. Neurosci Biobehav Rev 42: 180–192.

36. Langner R, Eickhoff SB (2013): Sustaining attention to simple tasks: a meta-analytic review of the neural mechanisms of vigilant attention. Psychol Bull 139: 870–900.

37. Mitko A, Rothlein D, Poole V, Robinson M, McGlinchey R, DeGutis J, et al. (2019): Individual differences in sustained attention are associated with cortical thickness. Hum Brain Mapp 40: 3243–3253.

38. Grasby KL, Jahanshad N, Painter JN, Colodro-conde L, Bralten J, Derrek P, et al. (2018): The genetic architecture of the human cerebral cortex.

39. Grotzinger AD, Mallard TT, Liu Z, Seidlitz J, Ge T, Smoller JW (2023): Multivariate genomic architecture of cortical thickness and surface area at multiple levels of analysis. Nat Commun 14: 946.

40. Marek S, Tervo-Clemmens B, Calabro FJ, Montez DF, Kay BP, Hatoum AS, et al. (2022): Reproducible brain-wide association studies require thousands of individuals. Nature 603: 654–660.

